# Unsupervised machine learning identifies distinct molecular and phenotypic ALS subtypes in post-mortem motor cortex and blood expression data

**DOI:** 10.1101/2023.04.21.23288942

**Authors:** Heather Marriott, Renata Kabiljo, Guy P Hunt, Ahmad Al Khleifat, Ashley Jones, Claire Troakes, Project MinE ALS Sequencing Consortium, TargetALS Sequencing Consortium, Abigail L Pfaff, John P Quinn, Sulev Koks, Richard J Dobson, Patrick Schwab, Ammar Al-Chalabi, Alfredo Iacoangeli

## Abstract

**Background:** Amyotrophic lateral sclerosis (ALS) displays considerable clinical, genetic and molecular heterogeneity. Machine learning approaches have shown potential to disentangle complex disease landscapes and they have been utilised for patient stratification in ALS. However, lack of independent validation in different populations and in pre-mortem tissue samples have greatly limited their use in clinical and research settings. We overcame such issues by performing a large-scale study of over 600 post-mortem brain and blood samples of people with ALS from four independent datasets from the UK, Italy, the Netherlands and the US.

**Methods:** Hierarchical clustering was performed on the 5000 most variably expressed autosomal genes identified from post-mortem motor cortex expression data of people with sporadic ALS from the KCL BrainBank (N=112). The molecular architectures of each cluster were investigated with gene enrichment, network and cell composition analysis. Methylation and genetic data were also used to assess if other omics measures differed between individuals. Validation of these clusters was achieved by applying linear discriminant analysis models based on the KCL BrainBank to the TargetALS US motor cortex (N=93), as well as Italian (N=15) and Dutch (N=397) blood expression datasets. Phenotype analysis was also performed to assess cluster-specific differences in clinical outcomes.

**Results:** We identified three molecular phenotypes, which reflect the proposed major mechanisms of ALS pathogenesis: synaptic and neuropeptide signalling, excitotoxicity and oxidative stress, and neuroinflammation. Known ALS risk genes were identified among the informative genes of each cluster, suggesting potential for genetic profiling of the molecular phenotypes. Cell types which are known to be associated with specific molecular phenotypes were found in higher proportions in those clusters. These molecular phenotypes were validated in independent motor cortex and blood datasets. Phenotype analysis identified distinct cluster-related outcomes associated with progression, survival and age of death. We developed a public webserver (https://alsgeclustering.er.kcl.ac.uk) that allows users to stratify samples with our model by uploading their expression data.

**Conclusions:** We have identified three molecular phenotypes, driven by different cell types, which reflect the proposed major mechanisms of ALS pathogenesis. Our results support the hypothesis of biological heterogeneity in ALS where different mechanisms underly ALS pathogenesis in a subgroup of patients that can be identified by a specific expression signature. These molecular phenotypes show potential for stratification of clinical trials, the development of biomarkers and personalised treatment approaches.

## BACKGROUND

Amyotrophic lateral sclerosis (ALS) is a fatal neurodegenerative disease which displays considerable genetic heterogeneity. Mutations in approximately 40 genes are known to be linked with ALS and can explain the majority of familial cases and approximately 20% of sporadic cases^1^ (SALS). However, a further 130 genes have been proposed to contribute to its risk or act as disease modifiers^2,3^. In approximately 90% of people with ALS, the disease is labelled as sporadic, without an apparent family history of the disease, with the remainder classed as familial^4^. ALS is also phenotypically variable, with differences in age and site of onset (spinal-innervated muscles vs bulbar), the balance of upper and lower motor neuron involvement, rate of disease progression, and the presence of cognitive or non-motor symptoms^5^. Furthermore, a multitude of molecular processes have been implicated in its pathogenesis, in part due to the vast number of causative and modifier genes associated with ALS that code for diverse cellular functions^6^. It is therefore plausible that there is no universal approach to the treatment of people with ALS, especially given that many therapeutic strategies target specific molecular pathways. For example, the protective action of Riluzole on motor neurons is proposed to be the result of a reduction in glutamate-mediated excitotoxicity^7^.

Machine learning (ML) approaches can be used to help us to understand the genetic and molecular complexity and heterogeneity of ALS, for example, by finding patterns in biological and clinical data that distinguish some groups of patients from the others. These subgroups can aid in identifying the best candidates for therapeutics which target specific biological processes. ML has previously been applied to brain expression data to stratify people with SALS into molecular subgroups^8–11^ and has led to valuable insights into the genomic heterogeneity of ALS. However, some of these studies integrated samples from different brain regions to generate clusters and characterise their molecular architectures^10–12^. This design would not reflect motor neuron-specific ALS pathogenesis. Other studies adopted a case-control framework^8,9,11^, which could lead to reduced power given the potential decoupling between mechanisms underlying risk and clinical presentation^13–15^. Furthermore, previous work has not been validated in independent datasets or in different populations and did not investigate whether molecular subtypes identified in post-mortem brains are reflected in other tissues available pre-mortem. Such factors have greatly limited the applicability and impact of these results. We therefore aimed to identify and validate molecular and phenotypic patterns across multiple independent datasets, tissue types and populations, to generate gene expression derived molecular subtypes of ALS that can be utilised for stratification in in the design and interpretation of future research and clinical studies.

## METHODS

A schematic overview of our study protocol is highlighted in Figure 1.

**Figure 1.**
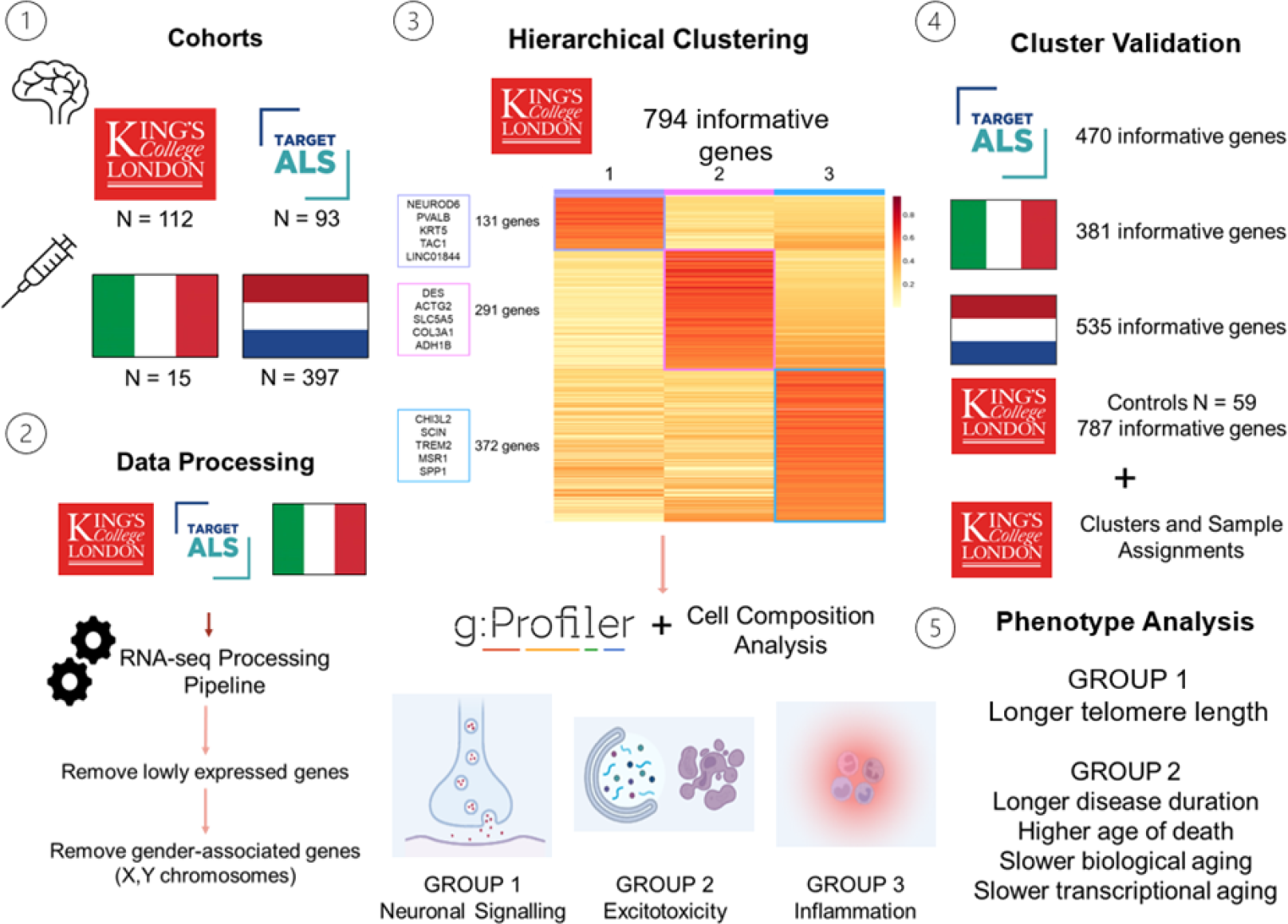
A graphical overview of our study design and analysis protocol. 1) A total of four cohorts were used in this study. Motor Cortex: KCL BrainBank (UK), cluster generation; TargetALS (USA), validation. Blood (PBMC): Zucca (Italy), validation; van Rheenen (Netherlands), validation. 2) The cohorts for which bulk RNAseq sequencing data was generated underwent processing using an in-house pipeline. 3) The non-smooth nonnegative matrix factorisation algorithm (nsNMF) was ran using the 5000 most variably expressed genes from KCL BrainBank as input, with k=3, 100 runs and 1000 iterations. There were 794 genes which were informative and uniquely assigned to one cluster. Gene enrichment and pathway analysis was performed for each cluster to obtain their molecular architectures, followed by cell composition analysis of the samples assigned to each cluster. We identified three genetically and molecularly distinct clusters which reflect previously investigated mechanisms of ALS pathogenesis. 4) Linear discriminant analysis (using sample cluster assignments from KCL BrainBank to train the model) was applied to the replication datasets, using the intersection of dataset-expressed genes and KCL BrainBank-derived informative genes. 5) Subgroup phenotype analysis was performed on all datasets, using various collected clinical and omics variables.

### Study Cohorts

We obtained raw post-mortem primary motor cortex bulk RNA sequencing data in FASTQ format from two datasets. The first, which was used to generate the clusters, consisted of 112 people from the UK with SALS from King’s College London and the MRC London Neurodegenerative Diseases Brain Bank (KCL BrainBank)^16^. We additionally obtained matching whole genome sequencing (WGS), methylation data and clinical data for the KCL BrainBank samples from Project MinE^16,17^ to perform subgroup clinical and omics-based phenotype analysis. For validation of KCL BrainBank-derived cluster expression signatures, 168 US samples from 93 people with SALS of North European ancestry, present in the Target ALS Human Post-mortem Tissue Core (TargetALS) were used. For further validation of KCL BrainBank-derived clusters, we also processed two peripheral blood mononuclear cell (PBMC) datasets; 1) bulk RNA sequencing data in FASTQ format of 15 Italian people with SALS (Zucca)^18^ (NCBI GEO Accession: GSE106443 and GSE115259), and 2) hg18-aligned log2 transformed and quantile normalised microarray gene probe intensities of 397 Dutch people with ALS (van Rheenen)^19^ (NCBI GEO Accession GSE112681). To determine if the clusters were ALS-specific, we also used RNA sequencing data from 59 healthy controls in the KCL BrainBank. Sequencing specific methods are described in more detail in the Supplementary Methods. The basic demographics of each of the datasets used in this study are detailed in Supplementary Table 1.

### Bulk RNA Sequencing Data Processing

Paired FASTQ files from KCL BrainBank, TargetALS and Zucca datasets were interleaved using BBMap reformat v38.18.0 under default options before adapters were right-clipped and both sides of each read were quality-trimmed with BBMap bbduk v38.18.0. The interleaved FASTQ files were aligned to hg38 using STAR v2.7.10a^20^ under default settings. Raw transcript counts for each gene were generated on a sample-wise basis before merging into dataset-specific matrices. The processing pipeline is available at https://github.com/rkabiljo/RNASeq_Genes_ERVs. Raw counts were normalised using the *estimateSizeFactors* function of DESeq2, before lowly expressed genes and sex chromosomes were removed. The whole dataset was standardized using the variance stabilising transformation *(vsd)* function in DESeq2^21^.

### Hierarchical Clustering of KCL Samples

Our hierarchical clustering was based on a protocol that was previously used to identify cortical molecular phenotypes of ALS^10^. Briefly, the 5000 most variably expressed genes, selected based on the highest median absolute deviation values, were extracted from the KCL BrainBank gene expression matrix. Unsupervised hierarchical clustering was then performed with the non-smooth negative factorisation (nsNMF) algorithm, using helper functions outlined in the SAKE package^22^. The optimal number of clusters was identified by running nsNMF with 100 runs and 1000 iterations for different values of k (2 to 10). Cluster estimation results are available in Supplementary Table 2. We then ran the nsNMF algorithm with k = 3, 100 runs and 1000 iterations, with the resulting consensus matrix showing a clear separation of samples (Supplementary Figure 1). Informative gene and sample assignment for each of the three clusters was then extracted. The list of informative genes for each cluster was then used to characterise their molecular phenotypes by performing gene enrichment analysis using the GProfiler2 R package^23^. Genes from the whole KCL expression matrix were used as a custom gene background. The default g:SCS algorithm was used to assess significant enrichment for several process and pathway categories in the following databases: Gene Ontology (Biological Process (GO:BP), Molecular Function (GO:MF) and Cellular Component (GO:CC)), Kyoto Encyclopaedia of Genes and Genomes (KEGG), Reactome, CORUM, TRANSFAC and miRTarBase. Additionally, MetaCore™ (available at https://portal.genego.com) was used to construct cluster-specific gene pathway networks using the *‘analyze network’* algorithm under default options, with the network that displayed the highest significance selected as the one that most defines the cluster.

### Cell Type Composition Analysis

Differences in cell composition between the samples in each cluster for both KCL BrainBank and TargetALS datasets were assessed with the BRETIGEA R package^24^ under default options for the following cell types: neurons, endothelial cells, astrocytes, microglia, oligodendrocytes and oligodendrocyte progenitor cells (OPCs). The singular value decomposition values, which gives us a measure of the relative contribution of each cell type to each cluster, were averaged for each cell type before differences in composition were calculated using ANCOVA with age at death and post-mortem delay included as covariates, with Bonferroni-corrected p-value of <0.05 denoting significance.

### Subgroup Phenotype Analysis

To reveal and compare the phenotypic architecture of each cluster, we extracted several clinical and omics variables from each dataset. Due to variations in the phenotypic information collected and accessibility of other omics data, we could not extract some phenotypic variables for all datasets. A breakdown of the collected phenotypic variables for each dataset is available in Supplementary Table 3. Transcriptional age acceleration was calculated by using RNAAgeCalc^25^ to obtain tissue-specific transcriptional age estimates for each dataset before being subtracted from the chronological age (age at death for KCL BrainBank and TargetALS, age at last blood draw for Zucca and van Rheenen). Telomere length and mitochondrial DNA copy number were obtained by applying TelSeq v0.0.2 ^26^ and fastMitoCalc v1.2^27^ to the whole genome sequencing BAM files, respectively. Biological age was estimated from the methylation beta-value matrix using CorticalClock^28^ before acceleration was calculated by subtracting each value from the age at death. Differences between clusters were assessed using one-way ANOVA, with post-hoc Tukey’s test used to determine subcluster-specific trends. The normality of each variable for each dataset was assessed using the Shapiro-Wilk test, with any variables that were non-normally distributed (p-value < 0.05) being log-transformed before analysing with one-way ANOVA. Results were corrected for sex. Additionally, we applied a Cox proportional-hazards model to assess differences in age of onset among clusters by combining samples from both KCL BrainBank and TargetALS datasets. A p-value of <0.05 denotes significance.

#### Code Availability

The implementation of our class assignment model based on the KCL BrainBank data, can be used to assign class membership to new expression samples (both microarray and RNAseq) and is publicly available at https://alsgeclustering.er.kcl.ac.uk. The code for the analyses performed in this study is available at https://github.com/KHP-Informatics/HierarchicalClusteringALS/.

## RESULTS

The nsNMF algorithm identified 794 of the 5000 most variably expressed genes as being the most informative for defining the clusters. Each informative gene was uniquely assigned to one cluster, yielding three genetically distinct clusters, each with a unique gene expression profile. There were 131, 291 and 372 genes which defined clusters 1, 2 and 3 respectively (Figure 2A). Further details of the genetic composition of each cluster are available in Supplementary Table 4. The larger proportion of the people with ALS (53.6%) were assigned to cluster 1, followed by cluster 2 (25%) and cluster 3 (21.4%), without substantial differences in male:female ratio (Figure 2B). Almost all C9 positive cases (87.5%) were assigned to cluster 1 (Table 1). Six known ALS-associated genes (*HSPB1, CAV1, CX3CR1, RNASE2, LUM, LIF*) were among the informative genes selected for the cluster signatures, with visible differences in their expression in samples assigned to their corresponding clusters (Supplementary Figure 2).

**Table 1.** Demographics and omics-based/clinical phenotypes for the samples assigned to each cluster for each dataset. NA represents values that could not be collected due to omics and clinical data availability.

|  | KCL BrainBank |  |  | TargetALS |  |  | Zucca |  |  | van Rheenen |  |  |
| --- | --- | --- | --- | --- | --- | --- | --- | --- | --- | --- | --- | --- |
|  | 1 | 2 | 3 | 1 | 2 | 3 | 1 | 2 | 3 | 1 | 2 | 3 |
| **Number of Samples (%)** | 60 (53.57) | 28 (25.00) | 24 (21.43) | 97 (57.7) | 28 (16.6) | 43 (25.6) | 13 (86.70) | 1 (6.65) | 1 (6.65) | 335 (84.38) | 33 (8.31) | 29 (7.31) |
| **Number of Samples with a posterior probability $\geq 80\%$ (%)** | NA | NA | NA | 88 (90.7) | 22 (78.6) | 31 (72.1) | 9 (69.2) | 1 (100) | 0 (0) | 275 (82.0) | 31 (72.1) | 11 (37.9) |
| **N Males: N Females (Ratio)** | 35:25 (1.4) | 15:13 (1.15) | 15:9 (1.67) | 60:37 (1.62) | 18:10 (1.80) | 21:22 (0.95) | 6:7 (0.86) | 0:1 (0) | 1:0 (0) | 205:130 (1.58) | 18:15 (1.20) | 16:13 (1.23) |
| **No. C9 positive individuals** | 7 | 1 | 0 | 11 | 0 | 4 | NA | NA | NA | NA | NA | NA |
| **Age at Symptom Onset in Years (mean ± SD)** | 59.2 ± 11.4 | 63.5 ± 13.8 | 59.7 ± 17.5 | 59.6 ± 11.1 | 64.9 ± 9.52 | 60.3 ± 11.5 | 63.6 ± 8.6 | 67.0 ± 0.0 | 65.0 ± 0.0 | 62.7 ± 11.9 | 57.9 ± 12.0 | 60.9 ± 12.3 |
| **Age at Blood Draw in Years (mean ± SD)** | NA | NA | NA | NA | NA | NA | 66.1 ± 9.8 | 69.0 ± 0.0 | 68.0 ± 0.0 | NA | NA | NA |
| **Age At Death in Years (mean ± SD)** | 62.5 ± 11.4 | 70.2 ± 11.4 | 64.2 ± 15.6 | 63.2 ± 10.2 | 69.5 ± 9.0 | 64.5 ± 8.9 | NA | NA | NA | NA | NA | NA |
| **Limb Onset (N)** | 36 | 10 | 17 | 65 | 22 | 17 | NA | NA | NA | NA | NA | NA |
| **Bulbar Onset (N)** | 15 | 7 | 5 | 14 | 5 | 21 | NA | NA | NA | NA | NA | NA |
| **Limb + Bulbar Onset (N)** | 1 | 1 | 0 | 7 | 0 | 1 | NA | NA | NA | NA | NA | NA |
| **Diagnostic Delay in Years (mean ± SD)** | 0.0015 ± 0.0013 | 0.00047 ± 0.00085 | 0.001 ± 0.0012 | 0.025 ± 0.32 | 0.073 ± 0.59 | 0.12 ± 0.35 | NA | NA | NA | NA | NA | NA |
| **Disease Duration in Years (median (IQR))** | 3.00 (1.96) | 1.71 (1.81) | 2.25 (1.75) | 3.00 (2.13) | 4.00 (3.48) | 2.00 (2.00) | NA | NA | NA | NA | NA | NA |
| **Post-mortem Delay in Hours (mean ± SD)** | 26.1 ± 12.10 | 26.0 ± 10.70 | 25.9 ± 13.90 | 9.9 ± 6.10 | 10.0 ± 7.45 | 12.0 ± 8.26 | NA | NA | NA | NA | NA | NA |
| **Mitochondrial DNA Copy Number (mean ± SD)** | 465 ± 22.0 | 457 ± 22.4 | 459 ± 17.3 | NA | NA | NA | NA | NA | NA | NA | NA | NA |
| **Telomere Length in Kilobytes (mean ± SD)** | 4.04 ± 0.46 | 3.98 ± 0.56 | 3.77 ± 0.42 | NA | NA | NA | NA | NA | NA | NA | NA | NA |
| **Transcriptional Age Acceleration in Years (mean ± SD)** | 6.16 ± 9.24 | 0.45 ± 10.90 | 5.59 ± 10.80 | 10.50 ± 8.63 | 4.19 ± 8.08 | 8.54 ± 8.44 | -23.50 ± 9.90 | -28.60 ± 0.00 | -26.93 ± 0.00 | -41.21 ± 11.66 | -36.68 ± 11.84 | -38.62 ± 11.69 |
| **Biological Age Acceleration in Years (mean ± SD)** | 5.99 ± 2.92 | 4.06 ± 4.65 | 7.93 ± 4.67 | NA | NA | NA | NA | NA | NA | NA | NA | NA |

**Figure 2.**
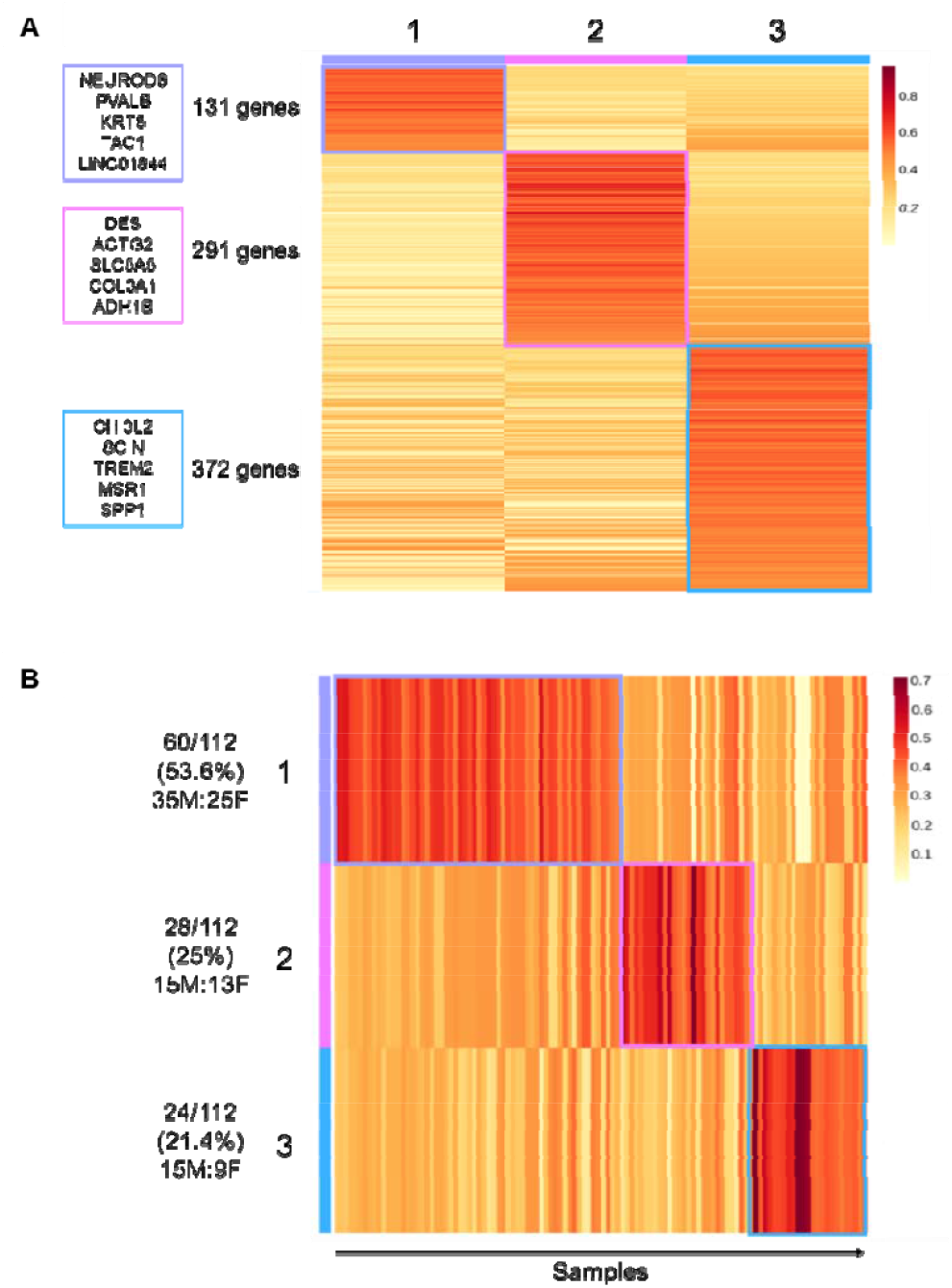
Informative gene and sample assignment for the KCL BrainBank, showing distinct separation of genes and samples to each cluster (1, 2, 3). A) Number of the 794 informative genes uniquely assigned to each cluster, with the top 5 contributing genes (defined by posterior probability) listed at the side. B) Distribution of cluster assignment of SALS cases alongside the male: female ratio. The coloured scale refers to the posterior probability value.

### Each cluster represents a molecularly distinct phenotype linked to ALS pathogenesis

Characterising the molecular architectures of each cluster by using gene enrichment and gene network analyses, we found that each cluster represents a distinct molecular phenotype. Cluster 1 was significantly enriched for various neuronal and synaptic signalling-related processes such as neuropeptide activity, cAMP signalling, and neuroactive ligand transcription, binding, and receptor interaction (Figure 3A, Supplementary Table 5). Network analysis revealed that a mitochondria specific signalling network is also present (Figure 3B, p= 1.05E-20). Led by *NXPH2, ATP12A, PTPRV, SV2C* and *C18orf42*, this network is enriched for mitochondrial ATP synthesis coupled electron transport and the aerobic electron transport chain.

**Figure 3.**
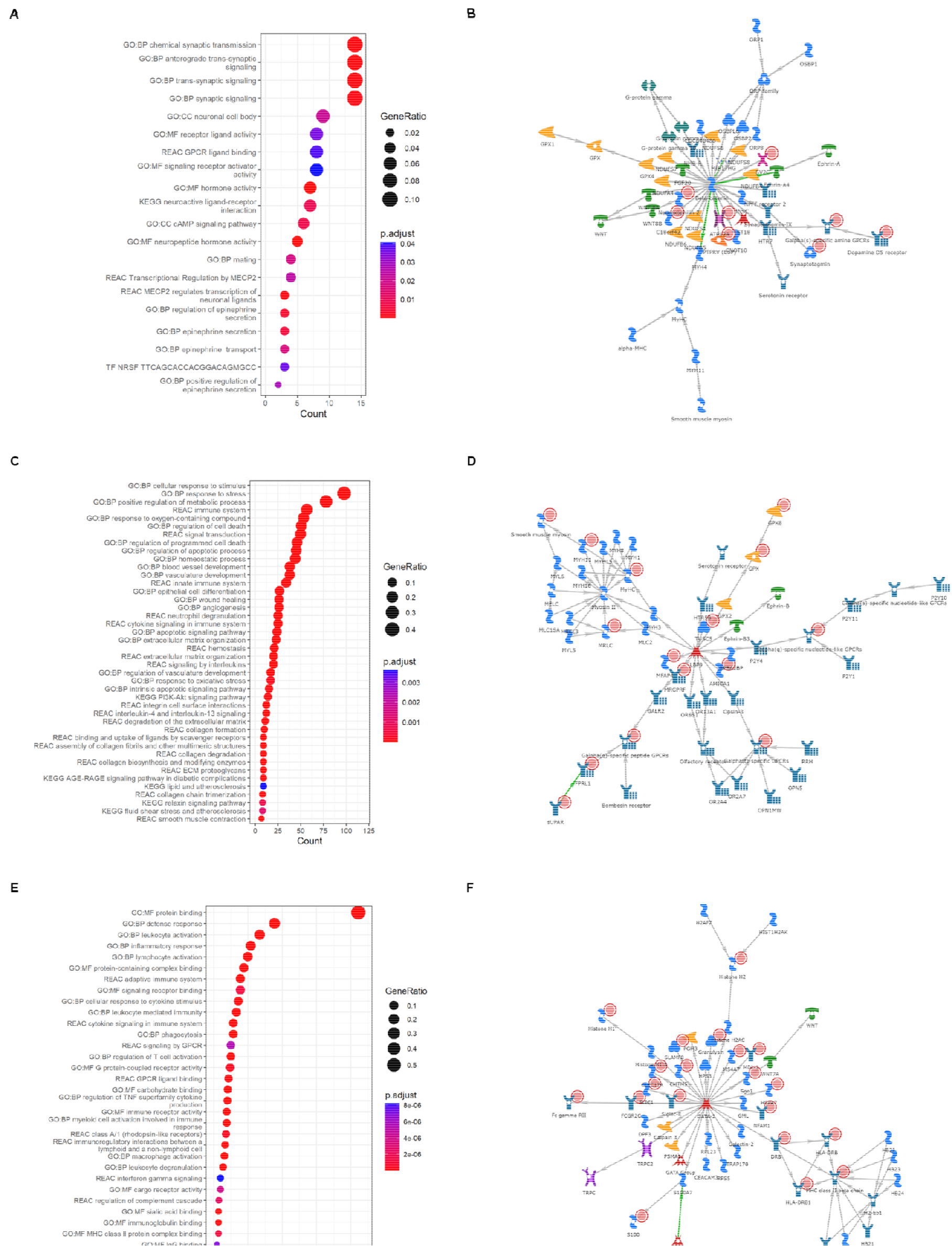
Results of gene enrichment and network analyses. Cluster 1: A) GProfiler2 reveals enrichment for various signalling-related processes. B) The most significant sub-cluster reveals a mitochondrial-specific signalling network. Cluster 2: C) GProfiler2 reveals enrichment for excitotoxicity, oxidative stress, anti-inflammatory and muscle system related processes. D) The most significant sub-cluster strengthens the support for muscle contraction processes being at the heart of this cluster. Cluster 3: E) GProfiler2 reveals enrichment for pro-inflammatory processes. F) The most significant sub-cluster reinforces the link to inflammation with the identification of an MHC Class 2 specific network. GO:BP – Gene Ontology Biological Process, GO:CC – Gene Ontology Cellular Components, GO:MF – Gene Ontology Molecular Function, KEGG: Kyoto Encyclopaedia of Genes and Genomes, REAC: Reactome, TF: Transfac. Red circles present in each network represent informative genes identified in each cluster. The descriptions of what the other symbols represent available in Supplementary Figure 3.

Cluster 2 was strongly linked with excitotoxicity, as shown by significant enrichments for oxidative stress, apoptotic signalling and cell death, and vasculature related processes such as angiogenesis, blood vessel development, epithelial cell differentiation and atherosclerosis (Figure 3C). Moreover, muscle-system and extracellular-matrix (ECM) specific enrichments (e.g., collagen synthesis and degradation, smooth muscle contraction, ECM proteoglycans and degradation) and anti-inflammatory pathways (interleukin-4 and interleukin-13 signalling, neutrophil degranulation) from Reactome were also associated with this cluster (Figure 3C). The muscle contraction theme was strengthened with GO:CC enrichments for banded collagen fibril, supramolecular fiber, myofibril, Z disc, I band, sarcomere and the actin cytoskeleton (Supplementary Table 6). Cluster 2 was also enriched for ALS-gene related NOS3-CAV1 CORUM complex (p = 0.018). Furthermore, the cluster 2 network (Figure 3D, p = 1.09E-17), which was driven by *MFAP4, FPRL1, TUSC5, MRGPRF* and *suPAR*, was associated with muscle contraction and actin-myosin filament sliding as well as phospholipase C-activating G protein coupled signalling. Cluster 3 represents a inflammatory phenotype, with biological process enrichment strongly associated with immune response in GO:BP and KEGG (Supplementary Table 7), as well as links with adaptive immunity, complement cascade and interferon gamma signalling in Reactome and immunoglobulin activity and major histocompatibility complex (MHC) class II in GO:MF (Figure 3E). Furthermore, C1q and TLR1-TLR2 CORUM complexes and viral diseases present in KEGG, such as Epstein-Barr disease, herpes simplex virus 1 and influenza A were among the most significant enrichments (Supplementary Table 7). Nine microRNAs were also significantly enriched in cluster 2 (including hsa-miR-335-5p, hsa-miR-146a-5p, hsa-mIR-124-3p, hsa-miR-29a-3p, and hsa-miR-204-5p), with hsa-miR-335-5p also being enriched in cluster 3 (Supplementary Tables 6 and 7). The cluster 3 network (Figure 3F, p = 1.47E-26), defined by *GNLY, HSPA7, SLAMF8, CLEC17A* and *Sgo1*, is MHC-class II specific and enriched for antigen processing, peptide antigen assembly, and presentation of peptides and polysaccharide antigens. Furthermore, the centre of the network, *GATA-2*, was the most significantly enriched TRANSFAC element in cluster 3 (*GATAD2A*, p = 9.56E-17, Supplementary Table 7).

### The identified molecular phenotypes replicate in independent post-mortem motor cortex data and blood datasets and are ALS-specific

To validate the KCL BrainBank derived clusters, we performed linear discriminant-driven cluster assignments of the TargetALS, Zucca and van Rheenen samples, using the intersection between the genes expressed in each one of them and the 794 genes that were used to define the clusters in the KCL BrainBank. 470, 381 and 535 were selected in this way for TargetALS, Zucca and van Rheenen datasets, respectively. In this analysis the linear discriminants were derived from the KCL BrainBank clustering. Samples from each dataset were assigned to one of the three clusters with high accuracy based on average posterior probability (diagonal cells in Figure 4A-C). A breakdown of the sample to cluster composition for all datasets is available in Table 1, with a visual inspection of their sample assignments available in Supplementary Figure 4. The posterior probability of assignment to each of the three clusters for each sample is available in Supplementary Table 8. To determine whether the clusters withheld validity in a control dataset, we applied the same modelling to the KCL BrainBank controls (demographics available in Supplementary Table 1). All controls were assigned to cluster 1 (Figure 4D). As the model is constrained to assign each sample to at least one class, we then sought to see if there were differences in the expression of the informative genes between cluster 1 cases and controls. We found that 66.4% of cluster 1 informative genes were significantly upregulated in cases that were assigned to cluster 1 (Supplementary Figure 5, Supplementary Table 9), supporting the ALS-specificity of the clusters.

**Figure 4.**
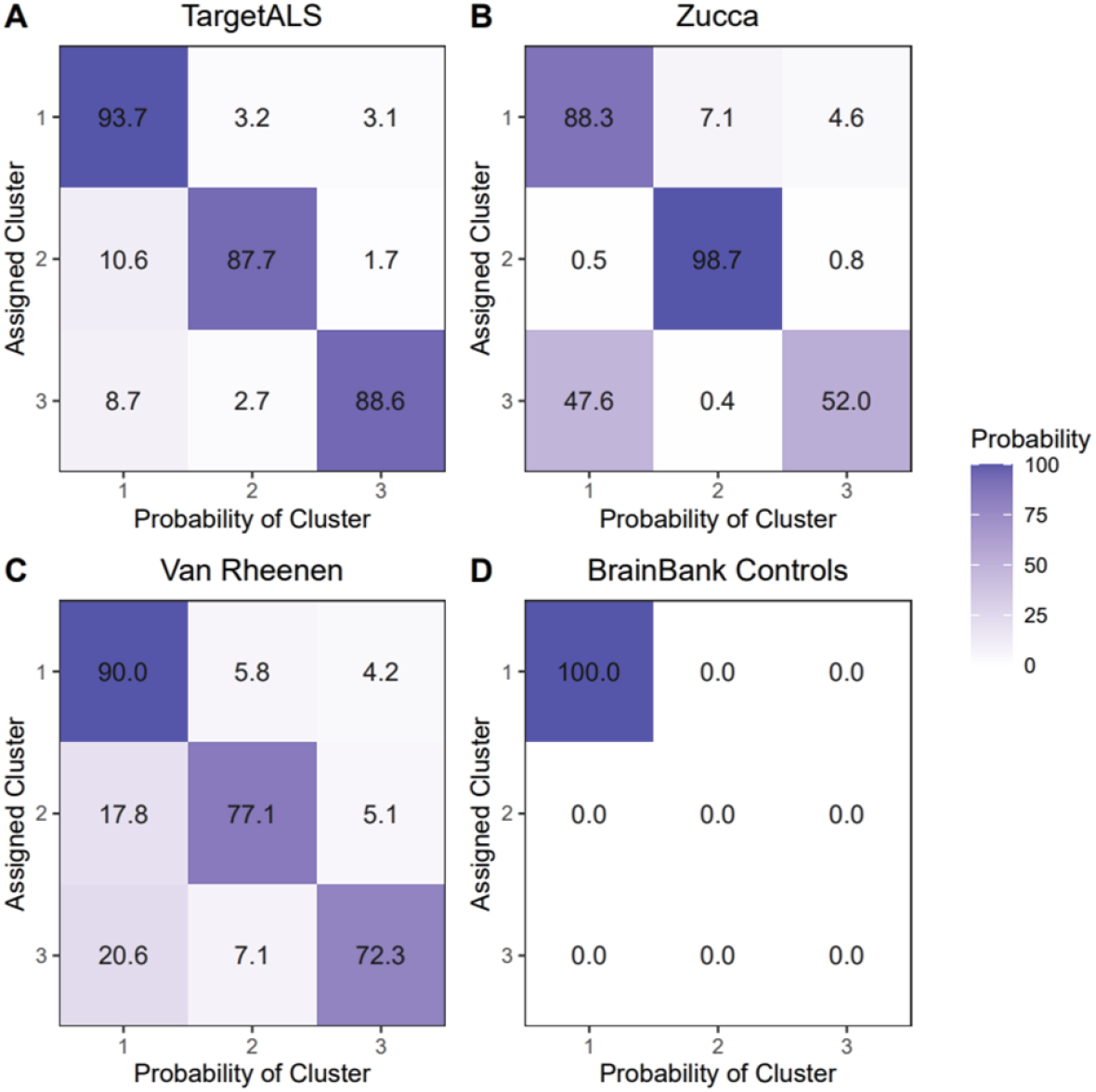
Results of linear discriminant analysis-based cluster assignment of A) TargetALS, B) Zucca, C) van Rheenen and D) BrainBank controls using the shared informative genes between each dataset and BrainBank cases and the BrainBank cases-defined cluster assignment to train the model. The x axis represents the average predicted posterior probability of being assigned to one of the three clusters, with the diagonals of the y axis representing the average posterior probability of being assigned to the correct cluster. Values are represented as percentages.

### Distinct cell types contribute to the molecular phenotypes

When performing cell deconvolution analysis for the KCL BrainBank and Target ALS datasets, we found that the samples that were assigned to each cluster had distinctive cell-type profiles which were very similar in both datasets (Figure 5). These profiles were reflective of the biological processes and networks identified during molecular phenotype analysis. Samples residing in cluster 1 had a higher neuronal cell contribution, whilst a higher endothelial cell composition was observed for cluster 2. Microglia were more prominent in cluster 3. Astrocytes, oligodendrocytes, and oligodendrocyte progenitor cells were also associated with cluster 3 in both datasets. The full results are available in Table 2.

**Table 2.** Statistical results of cell type contribution analysis using ANCOVA and Bonferroni post-hoc analysis to see cluster-specific trends. Results were corrected for sex and post-mortem delay.

|  | KCL BrainBank |  |  |  |  |
| --- | --- | --- | --- | --- | --- |
| Cell Type | Cluster 1 Mean ± SEM | Cluster 2 Mean ± SEM | Cluster 3 Mean ± SEM | ANCOVA (F-statistic, p value) | Post-Hoc Analysis (Bonferroni p-value) |
| Neurons | 0.036 ± 0.011 | -0.033 ± 0.016 | -0.052 ± 0.017 | 11.284, **3.0E-05** | 1 * 2; **0.0030**, 1 * 3; **1.4E-04**, 2 * 3; 1.0000 |
| Endothelial Cells | -0.053 ± 0.009 | 0.105 ± 0.013 | 0.009 ± 0.014 | 48.062, **1.28E-15** | 1 * 2; **4.98E-16**, 1 * 3; **0.001**, 2 * 3; **6.0E-6** |
| Microglia | -0.050 ± 0.010 | 0.023 ± 0.014 | 0.098 ± 0.015 | 36.140, **1.02E-12** | 1 * 2; **1.5E-04**, 1 * 3; **9.47E-13**, 2 * 3; **0.001** |
| Astrocytes | -0.053 ± 0.010 | 0.044 ± 0.015 | 0.080 ± 0.016 | 31.260, **2.04E-11** | 1 * 2; **1.0E-06**, 1 * 3; **3.05E-10**, 2 * 3; 0.304 |
| Oligodendrocytes | 0.005 ± 0.009 | -0.092 ± 0.014 | 0.095 ± 0.015 | 42.487, **2.62E-14** | 1 * 2; **2.82E-07**, 1 * 3; **4.0E-06**, 2 * 3; **9.67E-15** |
| Oligodendrocyte Progenitor Cells | -0.056 ± 0.010 | 0.056 ± 0.014 | 0.075 ± 0.015 | 36.101, **1.04E-12** | 1 * 2; **1.37E-08**, 1 * 3; **1.72E-10**, 2 * 3; 1.000 |
|  | TargetALS |  |  |  |  |
| Cell Type | Cluster 1 Mean ± SEM | Cluster 2 Mean ± SEM | Cluster 3 Mean ± SEM | ANCOVA (F-statistic, p value) | Post-Hoc Analysis (Bonferroni p-value) |
| Neurons | 0.270 ± 0.007 | -0.042 ± 0.013 | -0.034 ± 0.011 | 16.476, **3.25E-07** | 1 * 2; **3.9E-05**, 1 * 3; **2.2E-05**, 2 * 3; 1.000 |
| Endothelial Cells | -0.034 ± 0.007 | 0.056 ± 0.013 | 0.040 ± 0.010 | 28.611, **2.66E-11** | 1 * 2; **2.19E-08**, 1 * 3; **5.55E-08**, 2 * 3; 1.000 |
| Microglia | -0.024 ± 0.007 | -0.022 ± 0.013 | 0.075 ± 0.010 | 34.050, **5.52E-13** | 1 * 2; 1.000, 1 * 3; **8.99E-13**, 2 * 3; **6.03E-08** |
| Astrocytes | -0.035 ± 0.006 | -0.004 ± 0.011 | 0.085 ± 0.009 | 59.570, **6.43E-20** | 1 * 2; **0.047**, 1 * 3; **2.01E-20**, 2 * 3; **2.56E-08** |
| Oligodendrocytes | -0.022 ± 0.006 | -0.051 ± 0.011 | 0.086 ± 0.009 | 65.506, **2.40E-21** | 1 * 2; 0.071, 1 * 3; **1.79E-18**, 2 * 3; **2.52E-17** |
| Oligodendrocyte Progenitor Cells | -0.032 ± 0.007 | 0.026 ± 0.012 | 0.062 ± 0.010 | 31.812, **2.66E-12** | 1 * 2; **2.1E-04**, 1 * 3; **4.45E-12**, 2 * 3; 0.086 |

**Figure 5.**
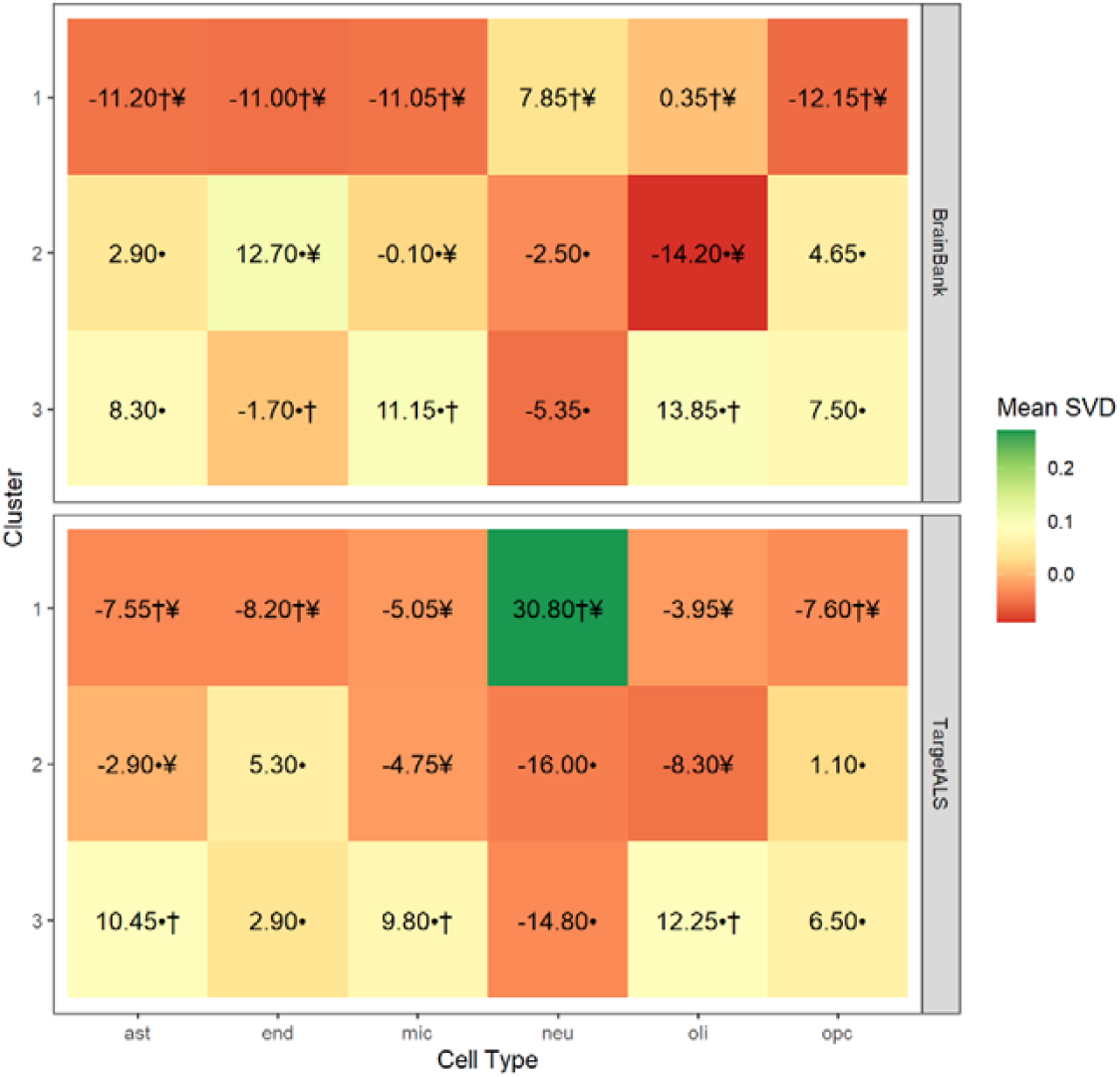
Cell type composition analysis in KCL BrainBank (top panel) and TargetALS (bottom panel) showing that samples in Clusters 1, 2 and 3 have a higher neuronal cell, endothelial cell, and microglia contribution, respectively, which reflects their enrichment for cell type-related processes. Astrocytes, oligodendrocytes, and oligodendrocyte progenitor cells are also associated with Cluster 3. The coloured scale corresponds to the mean singular value decomposition (SVD) of samples assigned to each cluster. Values represent the relative percentage change (Δ) of the mean SVD in that particular cluster compared to the mean SVD of the other two clusters for each cell type, with the symbols representing significant pairwise comparisons of the mean SVD in a particular cluster, compared to Cluster 1 (•), Cluster 2 (†) and Cluster 3 (¥). The cell types considered were neurons (neu), endothelial cells (end), astrocytes (ast), microglia (mic), oligodendrocytes (oli) and oligodendrocyte progenitor cells (opc).

### Clusters present different clinical outcomes and omics measures

In both KCL BrainBank and TargetALS, we observed that cluster 2 demonstrated differences in several phenotypic and omics measures (full results available in Table 3). For instance, cluster 2 compared to cluster 1, had a higher age of death (Figure 6A-B) and smaller transcriptional age acceleration (Figure 6C-D). This trend continues when looking at variables present in one of the two datasets, with a 3.87 year slower biological age acceleration being observed in cluster 2 compared to cluster 3 in KCL BrainBank (p-value 0.02), and a larger but albeit non-significant increase in disease duration in TargetALS samples assigned to cluster 2. We also found trends for longer telomere length and higher mitochondrial DNA copy number in cluster 1 in KCL BrainBank samples. When assessing differences in age of onset based on samples combined from KCL BrainBank and TargetALS, we found that samples residing in cluster 1 have a lower age of onset compared to clusters 2 and 3 (Figure 6E; p-value 0.013). For the Zucca and van Rheenen datasets, only age of onset and transcriptional age acceleration were available, for which there was no significant alteration in outcomes between clusters. The Zucca samples followed a similar trend of smaller transcriptional age acceleration in cluster 2 compared to cluster 1 (−5.1 years) and cluster 3 (−1.67 years) as the KCL BrainBank and TargetALS datasets, whereas the van Rheenen dataset seemed to follow the opposite trend (Table 3). This peculiarity continues when comparing differences in the age of onset of clusters in both brain and blood; the age of onset is higher in cluster 2 when looking at KCL BrainBank, TargetALS and Zucca datasets, with a lower age of onset in cluster 2 of van Rheenen compared to the other clusters (Table 3).

**Table 3.**
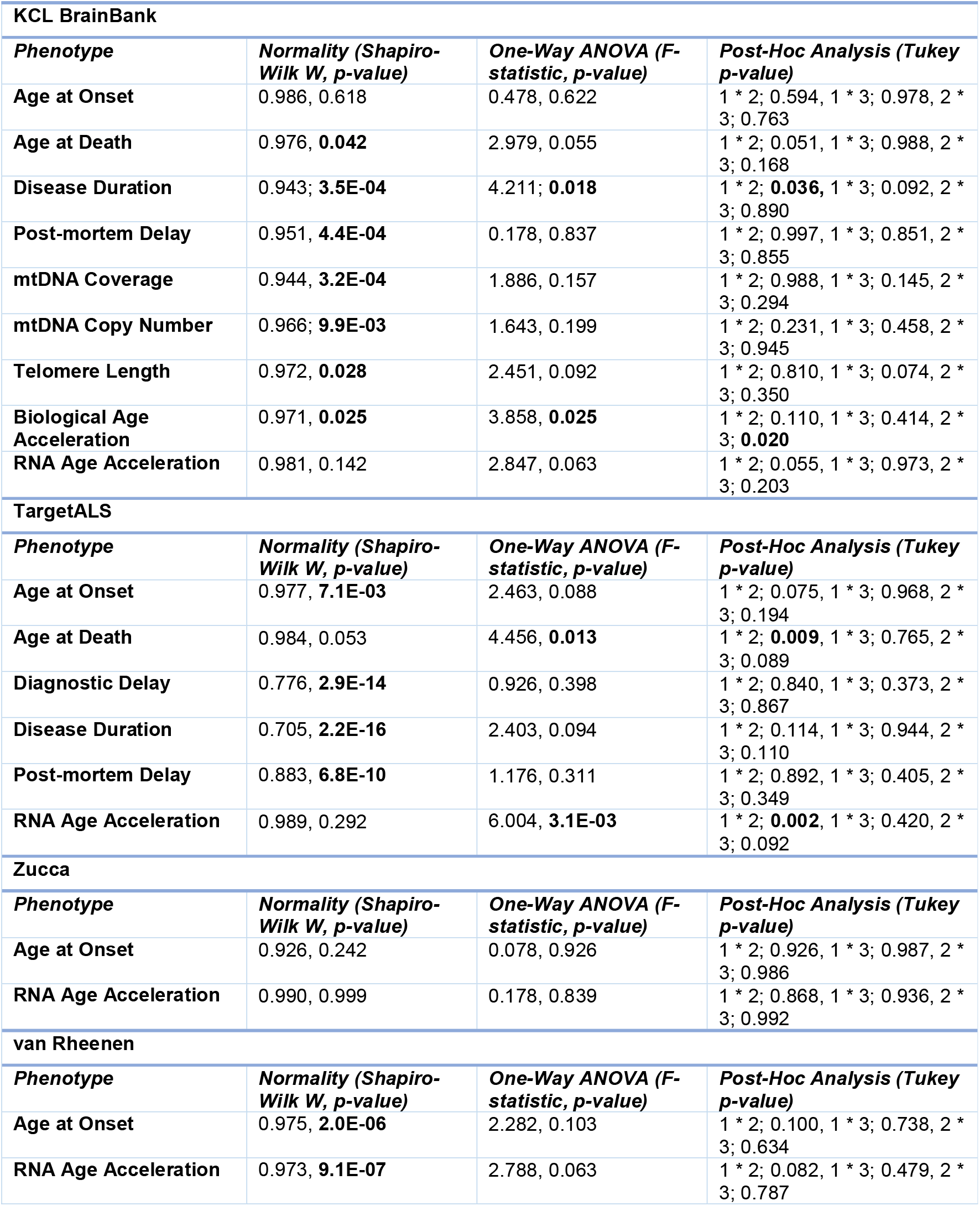
Statistical results of clinical and omics-based phenotype analysis. Variables that demonstrated non-normality via Shapiro Wilk were log transformed before running one-way ANOVA and post-hoc Tukey’s to assess cluster-specific trends. Results were corrected for sex.

**Figure 6.**
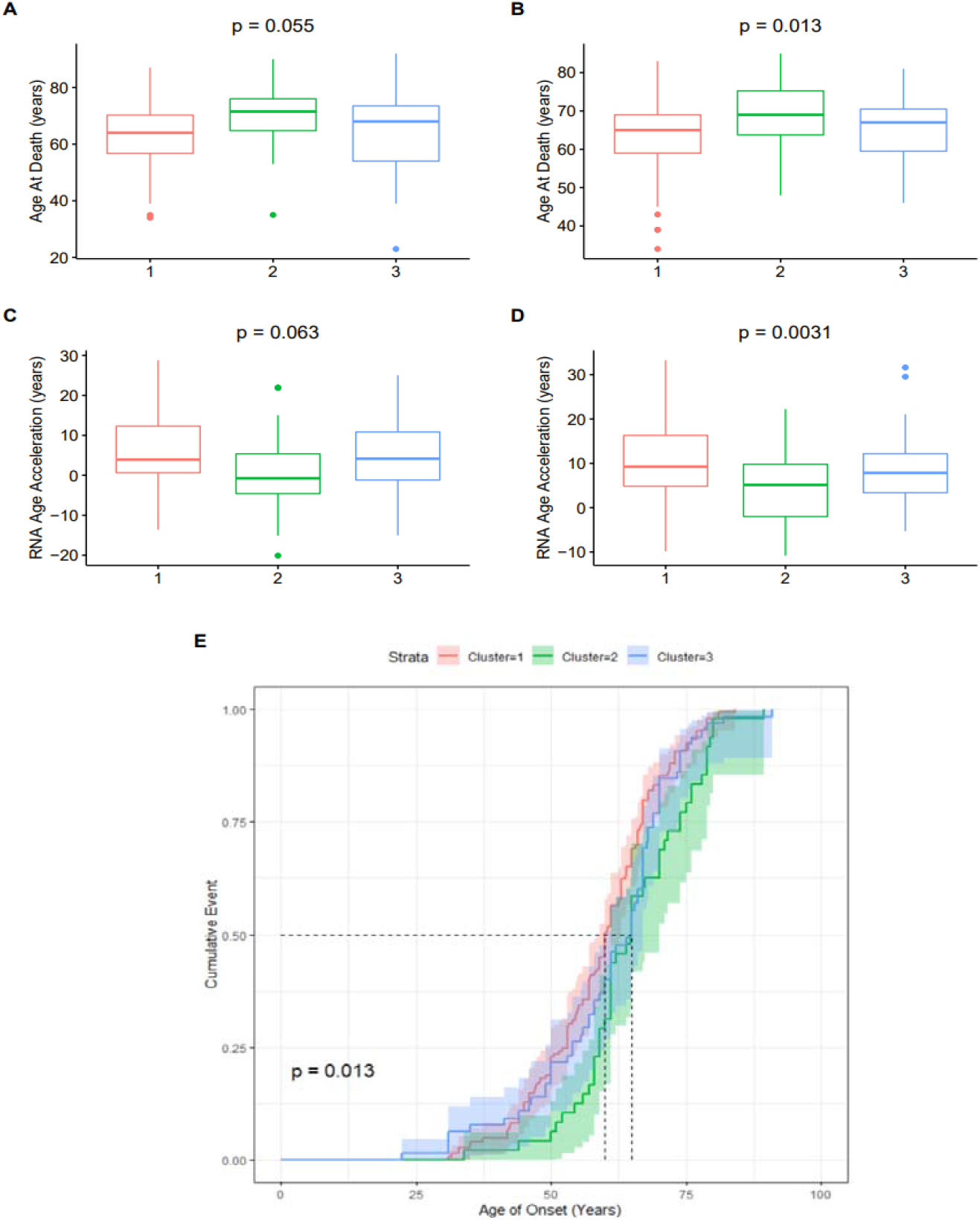
Subgroup phenotype analysis between samples residing in each cluster, comparing the age of death for A) KCL BrainBank and B) TargetALS, and transcriptional age acceleration for C) KCL BrainBank and D) TargetALS. P-values are from performing one-way ANOVA, corrected for sex. E) Cox proportional hazards model for the age of onset of samples from both BrainBank and TargetALS datasets, showing that samples from Cluster 1 have a significantly lower age of onset than Clusters 2 and 3.

## DISCUSSION

In this study, we used KCL BrainBank motor cortex gene expression data and machine learning to identify expression signatures which constitute three biologically homogeneous subgroups of SALS, which reflect three previously hypothesised key mechanisms of ALS pathogenesis. These molecular phenotypes were driven by different cell types, indicative of their main mode of pathogenesis. The mechanisms found in our study have been previously described using expression data^29^. In our study we showed that distinct mechanisms are relevant for distinct subgroups of patients and developed a model to stratify patient samples into these subgroups using post- and pre-mortem expression data. Moreover, our results were confirmed in independent motor cortex and blood tissue datasets from different populations of European ancestry.

### Cluster 1 – Neuronal Signalling Dysfunction

Genes which constitute the three main subgroups of cortical inhibitory GABAergic interneurons (*PVALB, SST, VIP*)^30^ were identified in this cluster, which is interesting given alterations in their excitability patterns cause global hyperexcitability of corticospinal neurons^31^. Hyperexcitability of motor cortex circuitry has long been hypothesised as a trigger for the spread of ALS pathology^32,33^. It is reasonable to propose that this hypothesis is supported by the evidence presented in this study, particularly given the cluster’s enrichment for anterograde trans-synaptic signalling. There were also several informative genes related to body mass index, metabolism and energy homeostasis (*LINC01844, ADCYAP1, CRH, CRHBP, CARTPT, VGF)*. These processes are linked with worse survival and progression outcomes in ALS^34–36^.

### Cluster 2 – Excitotoxicity / Muscle Contraction

Several muscle system related enrichments defined this cluster, which suggests that neuromuscular-based therapeutics could be beneficial for this subgroup of patients. This cluster was also enriched for anti-inflammatory signalling processes and contained several neuroprotective microglial secretory markers (*IL4R, TGFB1I1, TGFBI, CD163*)^37^ as well as the *MMP9* metalloproteinase gene, whose knockdown slows disease progression in ALS mutant models^38–40^. With microglia contributing minimally to this cluster, and better clinical and omics-based age outcomes defining the clusters phenotypic profile, we can postulate that a reversal of pro-inflammatory processes may be occurring in this SALS subpopulation. This is further supported by evidence that knockout of the ALS risk gene *CAV1*^41^ in endothelial cells, which were the drivers of pathogenesis for this molecular phenotype, can reduce innate immune system signalling via activation of endothelial nitric oxide synthase (NOS3)^42^; a complex of which was observed in our enrichment analysis.

### Cluster 3 - Inflammation

In this cluster, there was clear involvement of the major histocompatibility complex class II and the HLA complex (*HLA-DRA, HLA-DMB, HLA-DOA, HLA-DPA1, HLA-DRB1, HLA-DRB5, HLA-DRB6*), M1 or activated microglia (*CD14, CD86, TREM2, TYROBP, TMEM119, TMEM125)*^37^ and pro-inflammatory metalloproteinases (*MMP14*), as well as many immune related genes which were identified in other motor cortex and spinal cord SALS expression studies^8,43,44^. Three tentative ALS-related modifier genes (*LUM*^45^, *LIF*^46^, *CX3CR1*^47^), which are involved in proinflammatory processes^48–50^ and microglial-induced neuronal cell loss^51^, were also present in this cluster.

We also discovered that there were distinct clinical and omics-related outcomes that distinguished each cluster. Cluster 2 was associated with a slower progression and better outcome across both motor cortex and blood datasets. There are several plausible explanations as to why this trend was observed; the first is that more people assigned to this cluster may have a history of Riluzole usage than other clusters, as it modulates apoptosis, autophagy and excitotoxicity-related processes^52,53^. Another possibility is that there may be genomic variants present in inflammatory genes that abolish their effects. This theory is supported by the example of *IL18RAP*, which is an M1 secretory marker^37^ present in this cluster, of whom 3’UTR variants were recently found to protect against ALS, by impeding microglial-dependent motor neuron degeneration^54^. There is also evidence linking increased serum levels of the chronic inflammation marker suPAR, encoded by the informative gene *PLAUR*, with higher biological age acceleration in the normal population^55^. Therefore, suPAR could be a modulator of prognostic outcomes in SALS patients associated with this molecular phenotype. Telomere length was longer in cluster 1, which is also an important trend to investigate as there is mounting evidence supporting the association between longer telomere length and worsened severity of ALS^56,57^. Indeed, inhibition of the cluster-related gene LINC01844/miR-1255 can increase telomerase activity^58^, therefore the miR-1255 family should be studied as a potential biomarker of ALS.

Our analysis also revealed several known candidate gene biomarkers which could be exploited to stratify people with SALS. Cluster 3 contains several well studied serum and CSF biomarkers of ALS progression, such as *SPP1*^59^, the human chitinases *CHI3L1* and *CHI3L2*^60,61^, and complement C3^62^, in addition to prognostic and predictive CSF biomarkers such as *TREM2, LILRA2* and *ITGB2*^63^. Moreover, cluster 2 was enriched for several potential microRNA biomarkers. The most encouraging in terms of its impact on the molecular phenotype are miR-335-5p and miR-29b-3, as they are downregulated in ALS patients^64^. Additionally, their downregulation in model systems induces reactive oxygen species-mediated excitotoxicity^65^, and intrinsic apoptosis mediated motor neuron loss^66^.

There are several limitations of this study which will require further investigation in the context of our findings. First, only samples belonging to the KCL BrainBank dataset had matching multi-omics data, which meant that cluster-specific effects on omics variables could not be assessed in the other datasets. Likewise, both blood datasets had limited clinical information, which did not allow us to validate all possible clinical phenotype associations. Furthermore, the van Rheenen dataset displayed opposite trends in age-related outcomes. Some potential explanations are that microarray technology was used to obtain the transcriptomic profiles translating in a lower number of genes samples and lower class assignment accuracy, and that the Dutch population might present a more distinct structure compared to other European countries^67^. Furthermore, we did not integrate genomic variants into our analysis to further enhance our molecular classification, like recent studies that built upon their previous clustering analyses^11,68^.

In conclusion, we have demonstrated that people with ALS can be successfully stratified into molecularly and phenotypically distinct subgroups using gene expression data. Our results support the hypothesis that each mechanism underlies a distinct form of ALS pathogenesis and can be identified in patients via specific expression signatures. These molecular phenotypes discovered in a UK cohort, were validated in independent motor cortex and blood datasets, showing potential to be used for clinical trial stratification and the development of biomarkers and personalised treatments. We have developed a publicly available web app (https://alsgeclustering.er.kcl.ac.uk) to allow the broader scientific and clinical community to use our model for the stratification of samples and patients in their studies.

## Data Availability

All data produced in the present study are available upon reasonable request to the authors

## ACKNOWLEGEMENTS

H.M is supported by GlaxoSmithKline and the KCL funded centre for Doctoral Training (CDT) in Data-Driven Health. R.K receives funding from MND Scotland. G.P.H is supported by the Perron Institute for Neurological and Translational Science and the KCL funded centre for Doctoral Training (CDT) in Data-Driven Health. A.A.K is funded by ALS Association Milton Safenowitz Research Fellowship (grant number22-PDF-609.DOI :10.52546/pc.gr.150909.), The Motor Neurone Disease Association (MNDA) Fellowship (Al Khleifat/Oct21/975-799), The Darby Rimmer Foundation, and The NIHR Maudsley Biomedical Research Centre. J.Q is funded by The Darby Rimmer Foundation and the Motor Neurone Disease Association. S.K and A.L.P are funded by MSWA and the Perron Institute for Neurological and Translational Science. P.S is an employee and shareholder of GlaxoSmithKline plc. A.I is funded by the Motor Neurone Disease Association and The NIHR Maudsley Biomedical Research Centre. A.A-C is an NIHR Senior Investigator (NIHR202421) and has received support from an EU Joint Programme - Neurodegenerative Disease Research (JPND) project. The work is supported through the following funding organisations under the aegis of JPND - www.jpnd.eu *(United Kingdom, Medical Research Council* (MR/L501529/1; MR/R024804/1) *and Economic and Social Research Council* (ES/L008238/1)) and through the Motor Neurone Disease Association, My Name’5 Doddie Foundation, and Alan Davidson Foundation. The London Neurodegenerative Diseases Brain Bank at KCL has received funding from the MRC and through the Brains for Dementia Research project (jointly funded by Alzheimer’s Society and Alzheimer’s Research UK. We would like to acknowledge the Target ALS Human Postmortem Tissue Core, New York Genome Centre for Genomics of Neurodegenerative Disease, Amyotrophic Lateral Sclerosis Association and TOW Foundation for the RNA-sequencing data used in this publication. This study represents independent research part funded by the National Institute for Health Research (NIHR) Biomedical Research Centre at South London and Maudsley NHS Foundation Trust and King’s College London. The authors acknowledge use of the King’s Computational Research, Engineering and Technology Environment (CREATE) https://doi.org/10.18742/rnvf-m076. This work was supported by resources provided by the Pawsey Supercomputing Research Centre with funding from the Australian Government and the Government of Western Australia. The views expressed are those of the author(s) and not necessarily those of the NHS, the NIHR or the Department of Health and Social Care.

