## Supplementary materials for "Unsupervised machine learning identifies distinct molecular and phenotypic ALS subtypes in post-mortem motor cortex and blood expression data"

**SUPPLEMENTARY METHODS**

**Sequencing Details**

**RNA Sequencing**

**MRC London Neurodegenerative Diseases Brain Bank (KCL)**

The 100mg frozen tissue blocks were divided; one for RNA purification and the other for DNA. For each sample, a 30mg tissue block for RNA was homogenised using a Qiagen PowerLyzer 24 Homogenizer. Total RNA was purified from the homogenate using the standard protocol of the RNeasy Lipid Tissue Mini Kit (Qiagen), with on-column DNAse digestion. RNA integrity was estimated using Agilent Bioanalyzer 2100’s RNA 6000 Nano assays. RNA quantification was performed using a NanoDrop. Library preparation was performed using the standard Illumina TruSeq Stranded Total RNA Sample Preparation Guide with Ribo-Zero Human/Mouse/Rat (October 2013 Rev E.). Fragmentation steps were tailored to degrees of degraded RNA samples using Agilent Bioanalyzer 2100’s Nano assay results from the previous section. Libraries were validated using an Agilent Bioanalyzer 2100 to assess fragment size distribution. Library concentrations were estimated using a Qubit RNA High Sensitivity Assay Kit. Nanomolar (nM) concentrations were estimated using: nM = ng/ul X (1500/Average bp). Libraries were sequenced using Illumina HiSeq 4000 flow cells with 150bp paired-end reads with a target depth of 30 million clusters (60 million reads per sample).

**Target ALS Postmortem Tissue Core**

Information for the library preparation and RNA extraction of the TargetALS dataset is available at <http://www.targetals.org/wp-content/uploads/2020/11/README-1.zip>.

**PBMC Datasets**

Information for the library preparation and RNA extraction of the Zucca dataset is available at ^1^, and the microarray sequencing and normalisation protocol for the van Rheenen dataset is available at ^2^.

**Whole Genome Sequencing and Methylation Microarray Data**

For the KCL dataset, we also included matching whole genome sequencing and methylation microarray data collected under Datafreeze 2 of the Project MinE ALS sequencing consortium^3^. DNA was isolated from venous blood using standard methods. The DNA concentrations were set at 100 ng/uL as measured by a fluorimeter with the PicoGreen® dsDNA (Thermo Scientific, Waltham, MA) quantitation assay. DNA integrity was assessed using gel electrophoresis. The whole genome sequencing protocol is as follows: all samples were sequenced using Illumina’s FastTrack services (Illumina, San Diego, CA) on the Illumina HiSeq 2000 platform. Sequencing was 100 bp paired-end performed using polymerase chain reaction (PCR)-free library preparations and yielded ∼40x coverage across each sample. Binary sequence alignment/map formats (BAM) were generated for each individual. DNA methylation was analysed using Illumina Infinium EPIC array following the standard Infinium HD array methylation protocol (Illumina).

**SUPPLEMENTARY FIGURES**

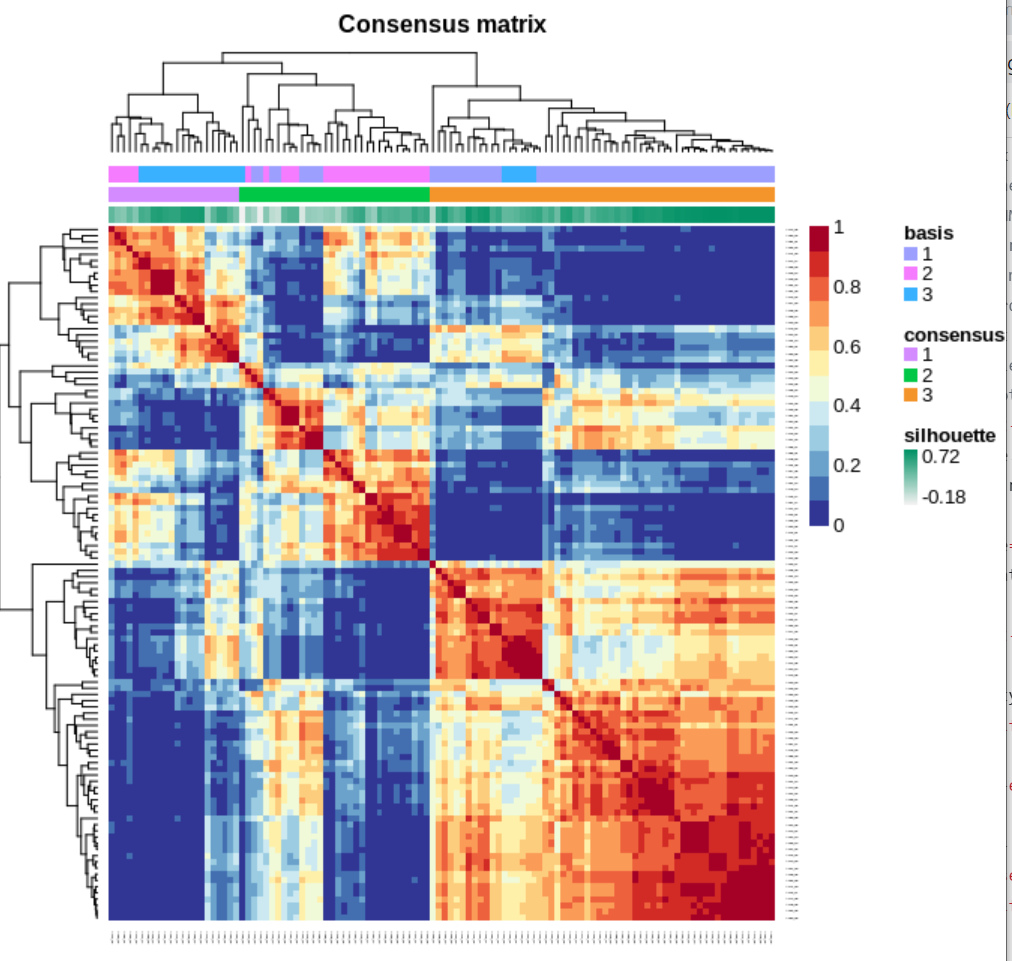

Supplementary Figure 1. Cluster construction with the KCL BrainBank consensus sample matrix for the KCL BrainBank dataset after running the nsNMF hierarchical clustering algorithm with k = 3, 100 runs and 1000 iterations.

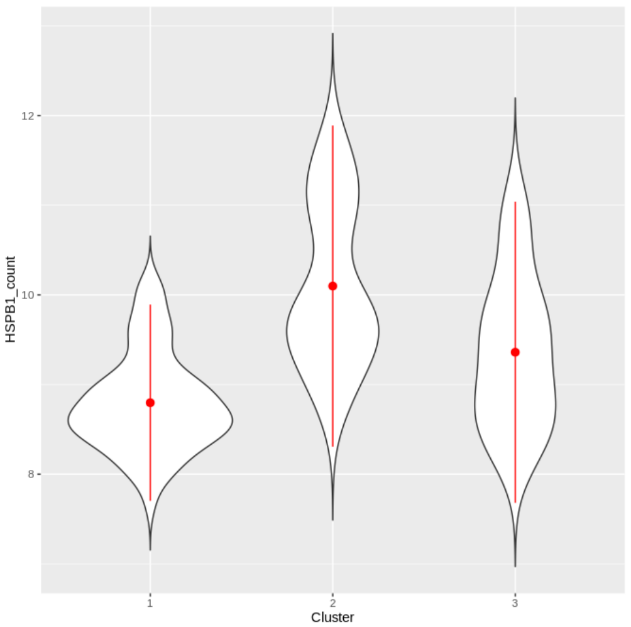

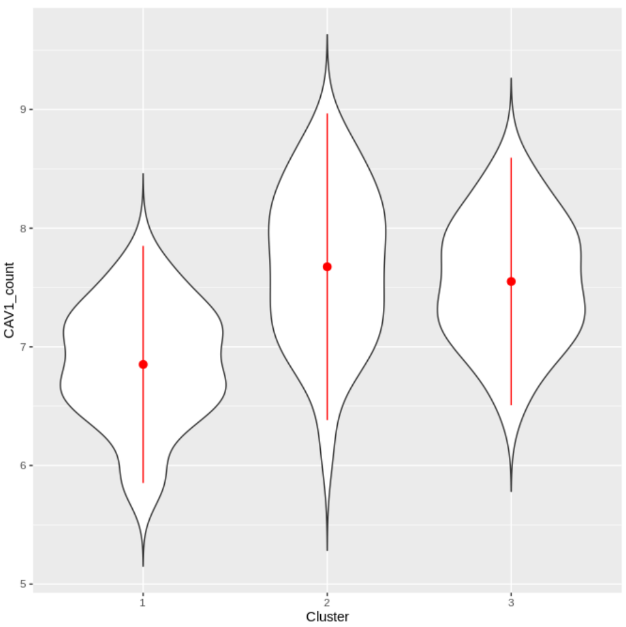

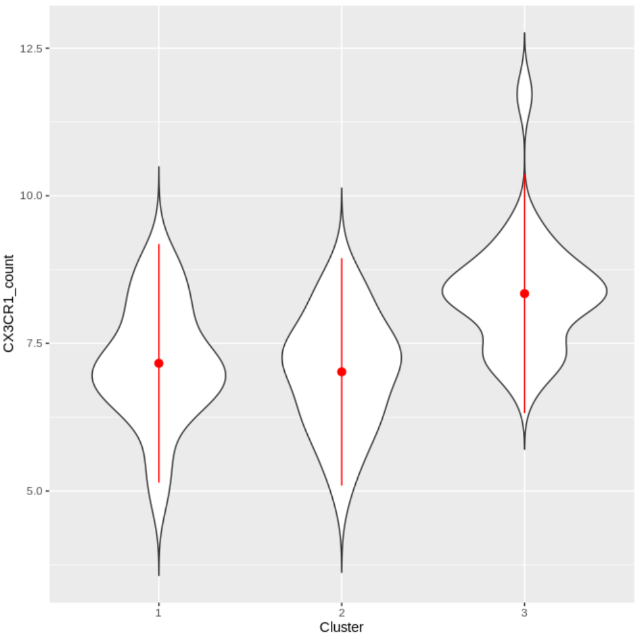

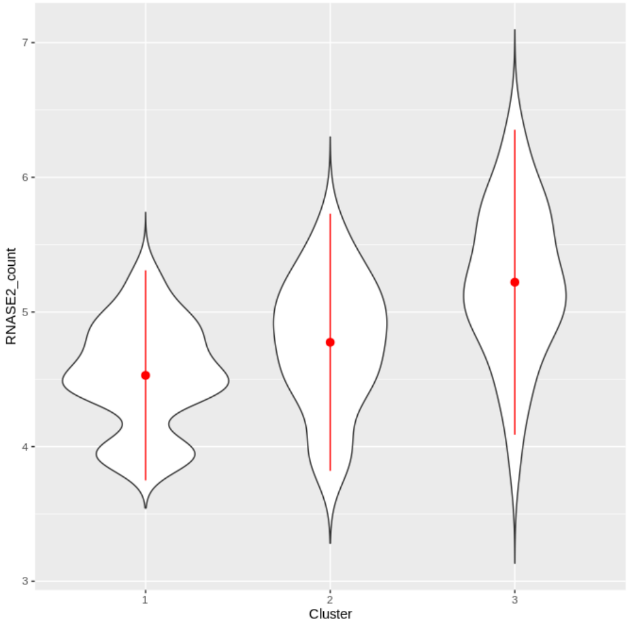

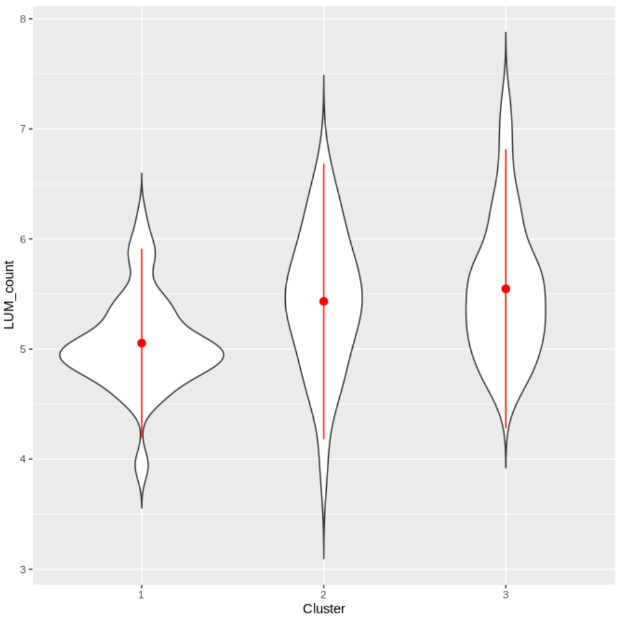

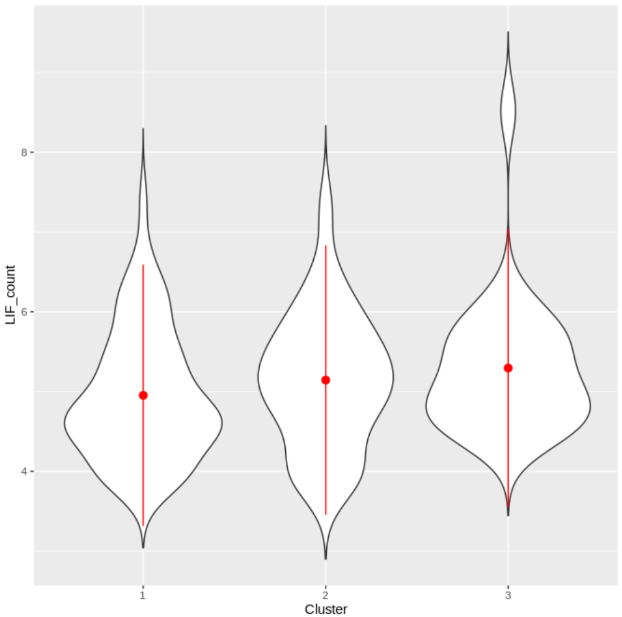

**A**

**B**

**C**

**D**

**E**

**F**

Supplementary Figure 2. Comparison of the expression of the six identified ALS-related genes A) HSPB1, B) CAV1, C) CX3CR1, D) RNASE2, E) LUM, and F) LIF, in each cluster, with the green asterisk representing the cluster that the gene is informative for.

*

*

*\

*\

*\

*\

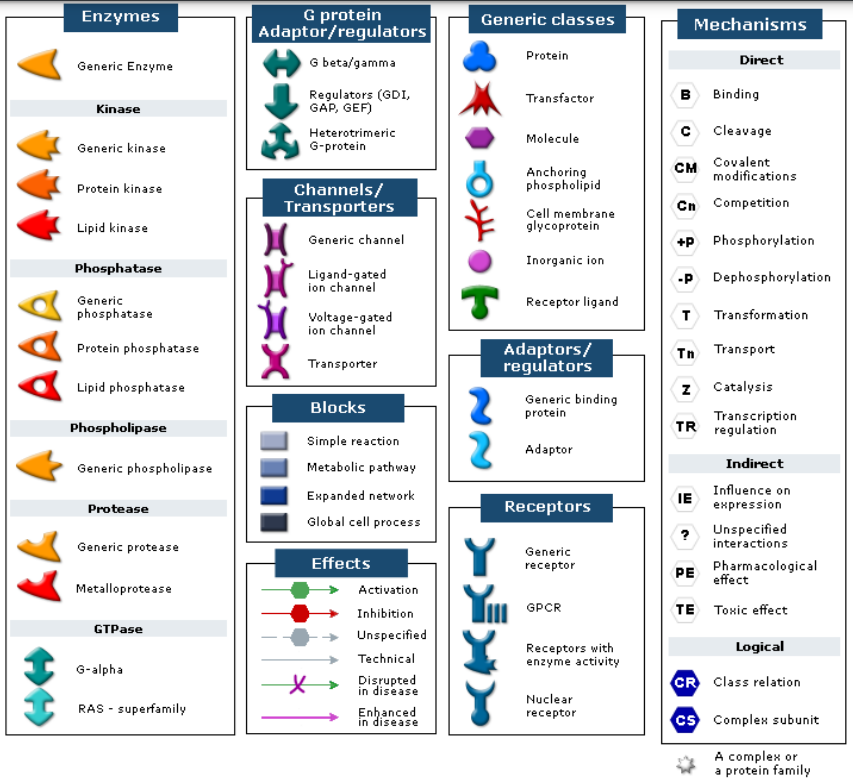

Supplementary Figure 3. A key of all of the symbols present in the sub-cluster networks.

**A**

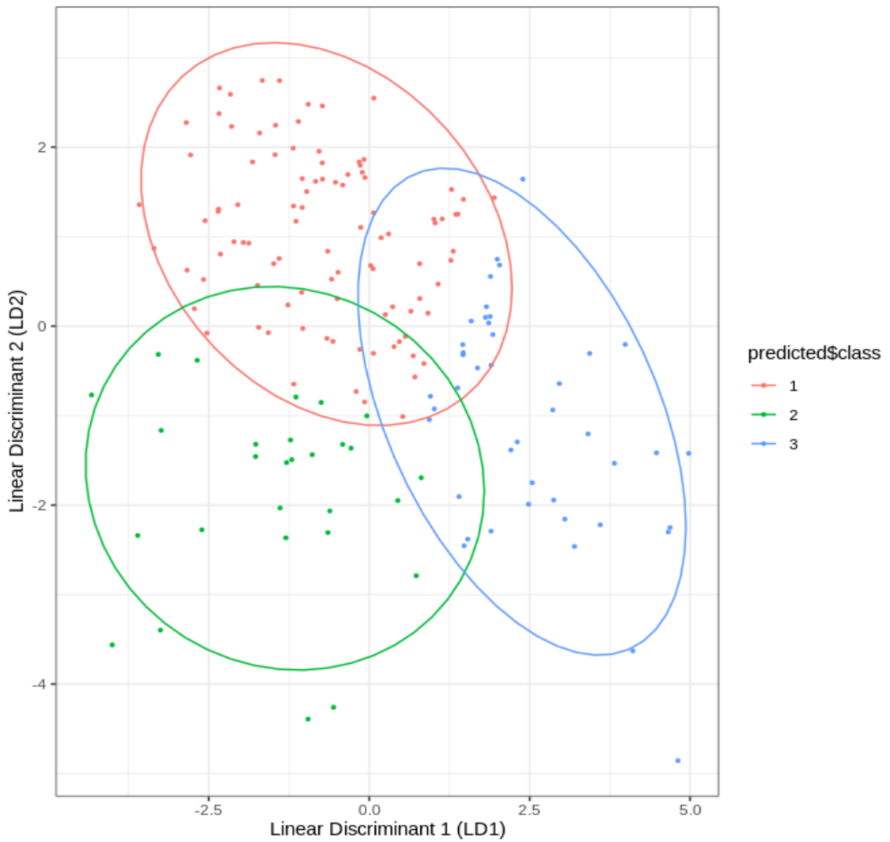

**B**

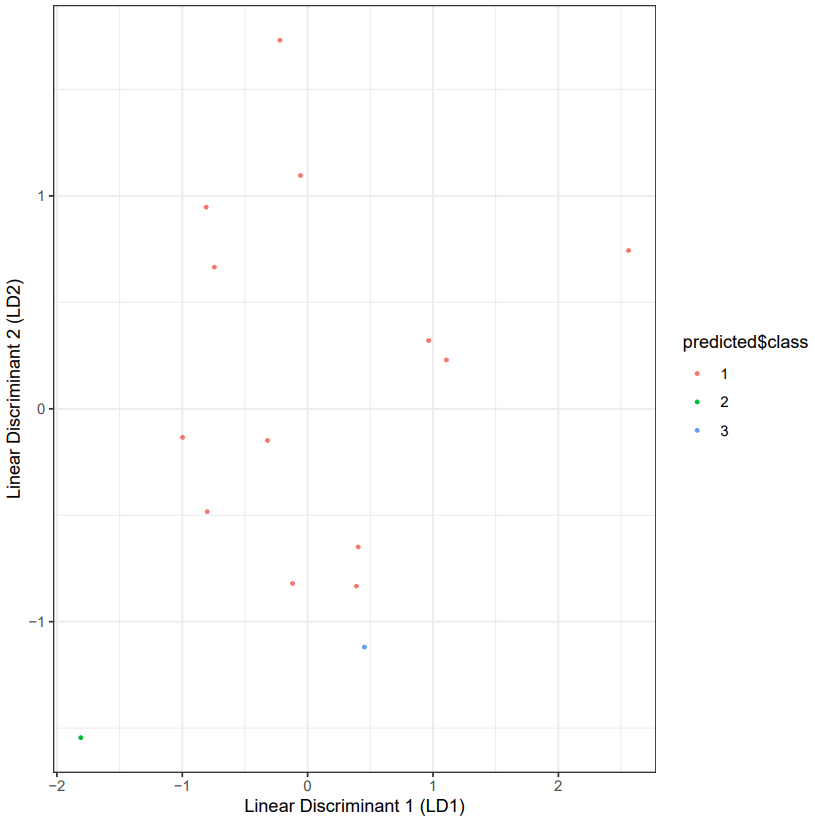

**C**

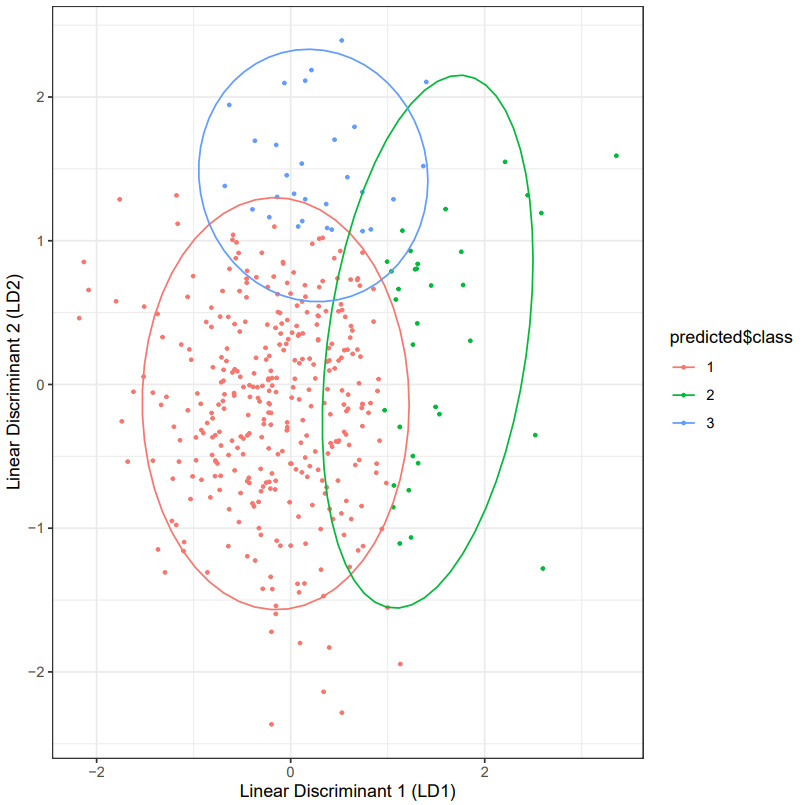

Supplementary Figure 4. Sample assignment of A)TargetALS, B) Zucca and C) van Rheenen datasets to the KCL BrainBank defined clusters. Linear discriminant analysis models were trained on the KCL sample assignments and informative genes shared with each dataset.

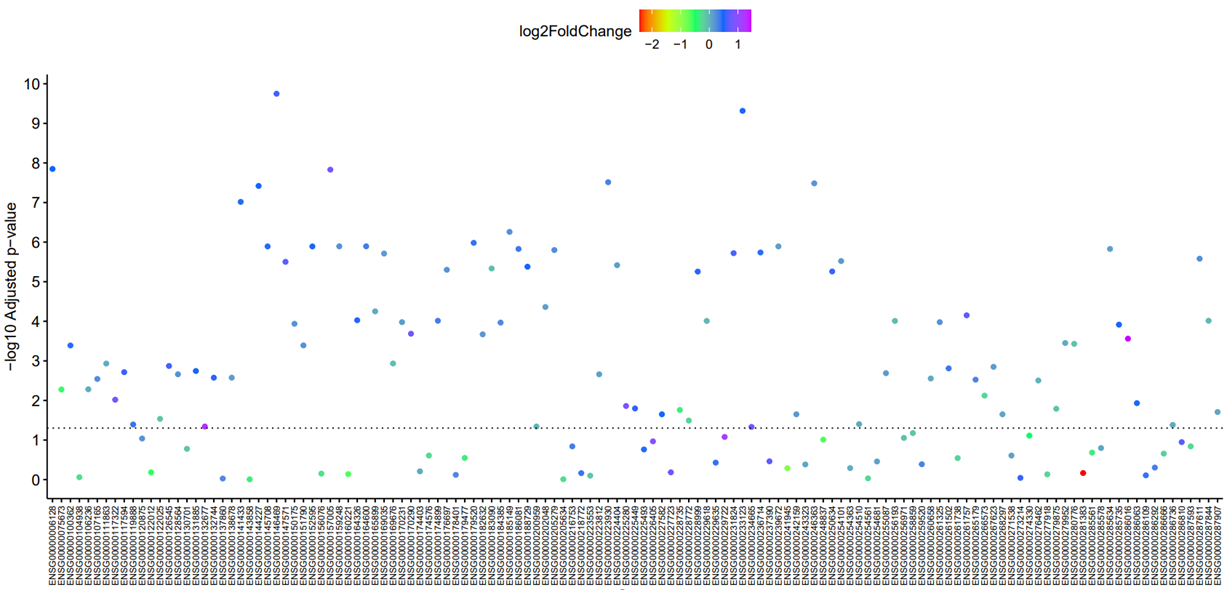

Supplementary Figure 5. Dotchart showing significant differences in expression of 87 of the 131 cluster 1 genes between KCL BrainBank cases and controls, coloured by log2 fold change. An increase in log2fold change means the gene is upregulated in cases, and vice versa. The dotted line represents the -log10 adjusted p-value that corresponds to a Benjamini-Hochberg p-value of 0.05.
